# MMA athletes demonstrate different brain vital sign profiles compared to matched controls at baseline

**DOI:** 10.1101/2024.03.26.24304733

**Authors:** Thayne A. Munce, Shaun D. Fickling, Shaquile Nijjer, Daniel N. Poel, Ryan C.N. D’Arcy

## Abstract

We investigated objective brain vital signs derived from event-related potentials (ERPs) for mixed martial arts (MMA) athletes and matched controls (N=24). Brain vital sign scans were acquired from 9 MMA athletes and 15 age- and sex-matched controls. Our analysis specifically compared differences in brain vital signs between MMA athletes and controls at baseline. We predicted that MMA athletes would show significant differences relative to controls due to their ongoing exposure to repetitive head impacts. Participants were scanned to extract three well-established ERPs: N100 for auditory sensation; P300 for basic attention; and N400 for cognitive processing. Scans were verified using automated reports, with N100, P300, and N400 amplitudes and latencies manually identified by a blinded reviewer. Groups were evaluated at the waveform level with a mass-univariate analysis using non-parametric resampling. Brain vital signs were compared across groups with a Kruskal-Wallis H-test for independent samples, with FDR correction for multiple comparisons. We identified significant differences between MMA athletes and controls. Specifically, there were significant N400 amplitude reductions, indicating that exposure to repetitive head impacts in MMA may be associated with changes in brain function.

## Introduction

Mixed martial arts (MMA) is a combat sport that continues to gain worldwide popularity. In competition, MMA athletes attempt to knock out, submit or out-point their opponent through a variety of striking, grappling and submission techniques. Through regular training and competition over many years, MMA athletes are routinely exposed to an undetermined amount of brain trauma, increasing their risk for concussion and other types of brain injury^1^.

Recent evidence has demonstrated that contact-sport athletes are not only threatened by concussions, but also subconcussive brain trauma associated with the frequency of repetitive head impacts^2,3^. While subconcussion is often imperceptible, as it does not have any recognizable clinical signs or symptoms, neurological and/or neuromuscular function may be impaired, which may limit performance and leave the athlete more susceptible to subsequent injury^4^. In the long term, exposure to repetitive, subconcussive head impacts has been linked to the onset of neurodegenerative disorders such as chronic traumatic encephalopathy (CTE) ^5,6^. Electroencephalography (EEG)-derived event-related potentials (ERPs)^7^, which represent brain responses to specific stimulus events, have increasingly been applied as objective, physiological measurements of cognitive function^8^. To translate this capability to the point-of-care, we developed and validated the brain vital signs framework^9^. The brain vital signs approach extracts three well-established target ERP responses: the N100 as a measure of auditory sensation^10^; the P300 as a measure of basic attention^10^; and the N400 as a measure of cognitive processing^11^. All three responses are elicited from a rapid auditory stimulation sequence comprised of randomly distributed auditory tones and spoken word pairs^9^. Each response is evaluated in terms of latency (milliseconds) and amplitude (microvolts) relative to standardized normative data and mapped as six metrics on a radar plot, where a symmetric hexagon shape represents a cognitive profile within the range of healthy norms. Together, the N100, P300 and N400 measurements of brain function provide enhanced sensitivity through objective neurophysiological measures to track cognitive changes in the brain.

Brain vital sign monitoring has recently been utilized as a sensitive measure for subconcussive impacts in contact sports. In an initial study by Fickling et al.^12^ of acute concussion in Junior A ice hockey players, an exploratory examination detected significant pre-versus post-season delay in N400 latency that was suggestive of delayed cognitive processing speed due to exposure to contact over the course of the season. These findings were subsequently replicated in two groups of ice hockey players (Junior A and Bantam) as well as in youth tackle football players^12-14^. These follow up studies demonstrated a significant linear relationship between brain vital signs changes and measures of head impact exposure. It was further shown that the changes in brain vital signs were significantly predictive of the total number of impacts that a player received, as measured by head-mounted accelerometers. In addition, brain vital signs changes were also significantly related to the total number of contact sport sessions (including games and practices) in which players participated. Collectively, these results, along with the emerging literature, indicate that exposure to repetitive subconcussive head impacts in a variety of contact sports is associated with measurable changes in brain function. However, it is unknown if similar subconcussive changes are concomitant with MMA participation.

## Objectives

The study objectives were to investigate subconcussive changes in brain vital signs in MMA athletes compared to matched controls at baseline. Our hypothesis predicted that MMA athletes would show significant differences in brain vital signs relative to controls due to their greater exposure to repetitive head impacts.

## Materials and Methods

### Participants

Overall, 34 participants were enrolled in the study, which was approved by Institutional review/ethics boards at Sanford Health and Advarra. There were 15 MMA athletes (N=15, Age=25.07±2.41, 2 female, 13 male) and 19 control participants (N=19, Age=26.21±3.11, 3 female, 16 male). Participants in the Control group were matched as closely as possible in relevant characteristics (e.g., age, sex, fitness level). Each participant provided written consent, according to the declaration of Helsinki. Inclusion criteria for participant recruitment was as follows: 1) MMA athletes: Adults (≥ 18 years of age) who were currently training (minimum of 3x/wk) in MMA and had been training for at least six months prior to study participation; 2) Control group: Age and sex matched adults (≥ 18 years of age) who were currently physically active (exercised for a minimum of 3x/wk at a moderate to high intensity).

The experimental design was a longitudinal, repeated-measures cohort study. Where applicable, baseline assessments were done at least 90 days from a previous fight (MMA group only) and 90 days (MMA) or 1 year (control) following medical clearance from a previously diagnosed concussion. While not the focus of the current analysis, the MMA group received up to three follow-up scans after a fight was completed.

Inconsistent fight schedules, participant compliance, and technical issues with data collection resulted in sample size attrition. Due to missing data and unequal samples, a within-subject, repeated-measures analysis was not possible. Accordingly, the primary analysis focused on between-group comparisons of MMA athletes and controls at baseline. There were 24 participants who successfully completed a baseline scan: 9 MMA athletes (N=9, Age=25.67±2.17, 0 female, 9 males) and 15 controls (N=15, Age=26.07±3.28, 3 female, 12 male). Two MMA athletes and four controls completed two baseline scans for purposes distinct from the current investigation. In cases of multiple baselines, analyses were completed using only the first successful scan.

### Brain vital signs data collection

Brain vital signs were extracted from ERPs using a g.Nautilus EEG cap (Gtec Medical Engineering, Austria) with embedded electrodes. After putting the cap on the participants head, g.GAMMAsys electrode gel was injected at each location for conductivity. A reference electrode was clipped to the right earlobe and disposable Ag/AgCl electrodes were used for ground (forehead), and electro-oculagram (EOG) recording from the supra-orbital ridge and outer canthus of the left eye. Skin-electrode impedances were maintained at <30k impedance at each site. A predefined auditory stimulus sequence was used^9^, in which acoustic stimuli (tones and word pairs) were delivered binaurally through earphones. Subjects were instructed to pay attention to the auditory stimuli while maintaining visual fixation on a cross located 2.0m away in a closed, quiet room. The same facility was used for all scans.

### Brain vital sign data processing

Raw EEG data were processed at the individual level using standard analysis methods. Data from three electrodes (Fz, Cz, & Pz) were sampled at 500Hz and bandpass filtered (0.1-20Hz). Ocular correction was applied using an adaptive filter^15^ with the EOG recordings as a reference. Denoised, filtered data were then segmented into epochs around common stimulus types (Epoch length: - 100ms pre-stimulus to +900ms post stimulus). Epochs were grand averaged across all electrodes to generate a single composite evoked potential waveform for each stimulus type (standard tone, deviant tone, congruent word, incongruent word). For each brain vital sign, response size (amplitude in microvolts) and timing (latency in milliseconds) were manually verified and recorded by an experienced blinded reviewer^16^. N100 and P300 were labelled on the deviant tone response, and the N400 was labelled on the incongruent word response.

### Statistical comparisons

Statistical analyses consisted of a time-series analysis of the amplitudes and latencies for the N100, P300, and N400. A mass-univariate analysis was conducted using non-parametric resampling to account for sample attrition issues. Group comparisons used a Kruskal-Wallis H-test for independent samples, with Benjamini-Hochberg Method of False Discovery Rate correction for multiple comparisons. We compared MMA athletes versus controls at baseline.

### Radar plot comparison

Standardized radar plots were generated to visualize multivariate changes in brain vital signs (N100, P300, and N400 responses) For any given response, larger amplitudes and faster (i.e. smaller) latencies are plotted on the peripheral of the figure. Normative (i.e. group median) results converge on a symmetric hexagonal profile. This method allows for all six metrics to be plotted radially on the same scale, and to visualize multivariate change either within or between groups.

## Results

Figure 1A depicts grand-average (top) and representative individual (bottom) waveforms showing the N100, P300, and N400 for the baseline comparison. Examination of the waveform results revealed a general decrease in all response amplitudes for MMA athletes relative to controls.

**Figure 1:**
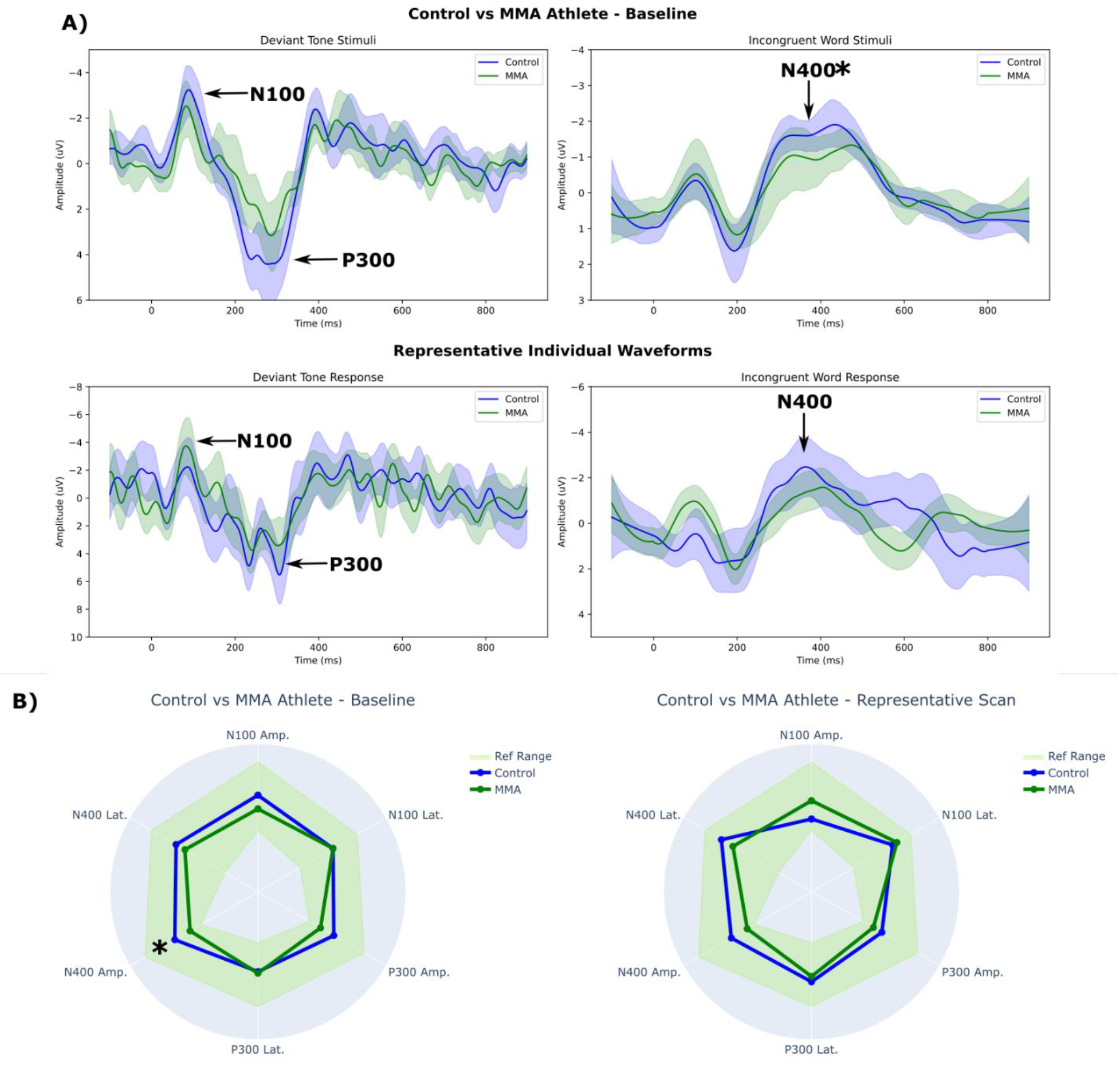
A) Waveform and B) Radar plot comparison between MMA athletes and controls at baseline. * p < 0.05 (corrected)

Figure 1B provides the radar plot brain vital sign comparison at the grand-average (top) and representative individual (bottom) levels. Brain vital sign profiles confirmed that while the reduction in response amplitude and N400 latency delay were present, only the reduced N400 amplitude was significant given the relatively high variability within the restricted sample size. Table 1 provides uncorrected and corrected P-values for each Brain Vital Sign.

**Table 1.** Brain vital sign measures for MMA athletes and controls across comparisons.

|  |  | h | P (uncorrected) | p (FDR corrected) |
| --- | --- | --- | --- | --- |
| Baseline Comparison | N100 amplitude | 1.4325 | 0.2314 | 0.3470 |
|  | N100 latency | 1.5172 | 0.2180 | 0.3470 |
|  | P300 amplitude | 2.1342 | 0.1440 | 0.3470 |
|  | P300 latency | 0.1507 | 0.6978 | 0.6978 |
|  | N400 amplitude | 7.0440 | 0.0080 | **0.0477\*** |
|  | N400 latency | 0.9680 | 0.3252 | 0.3902 |

## Discussion

The study investigated subconcussive changes in brain vital signs in MMA athletes compared to matched controls at baseline. The findings supported the hypothesis that MMA athletes would show significant brain vital sign differences relative to controls, presumably due to routine exposure to repetitive head impacts while participating in their sport.

Examination of the waveform differences between MMA athletes and controls showed a common reduction in N100, P300, and N400 response amplitudes. The current findings contribute to the growing body of literature that supports the link between repetitive head impacts and subconcussive impairment,. Given our recent ice hockey study (Breuer et al., 2023, unpublished), which demonstrated no detectable subconcussive impairments with specific daily dietary supplementation, future work should investigate potentially effective and accessible intervention options.

The current study represents another replication in support of the relationship between head impact exposure and brain vital signs^12-14^ (& Breuer et al., 2023, unpublished). Specifically, subconcussive changes in the cognitive N400 response have repeatedly been observed across all studies to-date. The replications have been across different contact sports (i.e., ice hockey, football, and mixed martial arts) as well as different age ranges (i.e., approximately ages 12-30 years old) in male athletes. It is worth noting that the particular N400 changes have varied in terms of latency delays, amplitude reductions, or both across the noted studies. The two factors are interdependent and interactive, with relative peak timing delays and response reductions representing a relative impairment in cognitive processing, specifically semantic processing^17-19^. On-going studies have further confirmed the N400 effects compared to non-contact control comparisons, across different sports, and between females and males (McCarthy et al., unpublished), with additional N100 and P300 changes being similarly detectable.

## Limitations

Our available subject pool of MMA athletes was limited, leading to a relatively low sample size. Moreover, we had to exclude some acquired EEG scans due to data quality, precluding us from conducting a planned longitudinal analysis. While subjects in the control group were not participating in contact sports during the study, other lifetime exposures were not accounted for that may have impacted our findings. Furthermore, we cannot differentiate if the measured baseline differences reflect an association with MMA participation in general, or the acute effects of recent MMA activity. Future studies with larger sample sizes will make it possible to evaluate specific factors related to MMA participation, such as fight history, outcomes and training methods that may have influenced our results. Moreover, future studies would benefit from incorporating fight performance metrics or using impact sensors to derive specific head impact measures of exposure. Likewise, a more robust longitudinal design would allow for direct assessment of brain vital sign changes over time in relation to MMA exposure measures.

## Conclusion

MMA athletes demonstrate significant brain vital sign differences in baseline scans compared to controls. Specifically, significant brain vital sign differences were detected as reduced amplitudes for the N400, with additional reductions observed in the N100 and P300. The findings indicate that exposure to repetitive head impacts in MMA may be associated with subconcussive changes in brain function.

### Transparency, Rigor, and Reproducibility Statement

For details, please refer to the Methods and Limitations sections.

## Data Availability

All data produced in the present study are available upon reasonable request to the authors

## Acknowledgements

The authors would like to thank the participants for their voluntary involvement in this research.

## Authorship Confirmation/Contributions

**Shaun Fickling:** Data Curation, Formal Analysis, Investigation, Visualization, Writing – Original Draft. **Shaquile Nijjer:** Visualization, Writing - Review and Editing. **Daniel Poel:** Investigation, Data Curation, Writing – Review and Editing. **Thayne Munce:** Conceptualization, Funding Acquisition, Methodology, Writing – Review & Editing. Supervision. **Ryan D’Arcy:** Conceptualization, Methodology, Supervision, Writing – Review and Editing.

